# The Impact of COVID-19 on NO_2_ and PM_2.5_ Levels and Their Associations with Human Mobility Patterns in Singapore

**DOI:** 10.1101/2021.12.02.21267165

**Authors:** Yangyang Li, Yihan Zhu, Jia Yu Karen Tan, Hoong Chen Teo, Andrea Law, Dezhan Qu, Wei Luo

## Abstract

The decline in NO_2_ and PM_2.5_ pollutant levels were observed during COVID-19 around the world, especially during lockdowns. Previous studies explained such observed decline with the decrease in human mobility, whilst overlooking the meteorological changes (e.g., rainfall, wind speed) that could mediate air pollution level simultaneously. This pitfall could potentially lead to over-or under-estimation of the effect of COVID-19 on air pollution. Consequently, this study aims to re-evaluate the impact of COVID-19 on NO_2_ and PM_2.5_ pollutant level in Singapore, by incorporating the effect of meteorological parameters in predicting NO_2_ and PM_2.5_ baseline in 2020 using machine learning methods. The results found that NO_2_ and PM_2.5_ declined by a maximum of 38% and 36%, respectively, during lockdown period. As two proxies for change in human mobility, taxi availability and carpark availability were found to increase and decrease by a maximum of 12.6% and 9.8%, respectively, in 2020 from 2019 during lockdown. To investigate how human mobility influenced air pollutant level, two correlation analyses were conducted: one between PM_2.5_ and carpark availability changes at regional scale and the other between NO_2_ and taxi availability changes at a spatial resolution of 0.01°. The NO_2_ variation was found to be more associated with the change in human mobility, with the correlation coefficients vary spatially across Singapore. A cluster of stronger correlations were found in the South and East Coast of Singapore. Contrarily, PM_2.5_ and carpark availability had a weak correlation, which could be due to the limit of regional analyses. Drawing to the wider context, the high association between human mobility and NO_2_ in the South and East Coast area can provide insights into future NO_2_ reduction policy in Singapore.

**Graphical Abstract:** 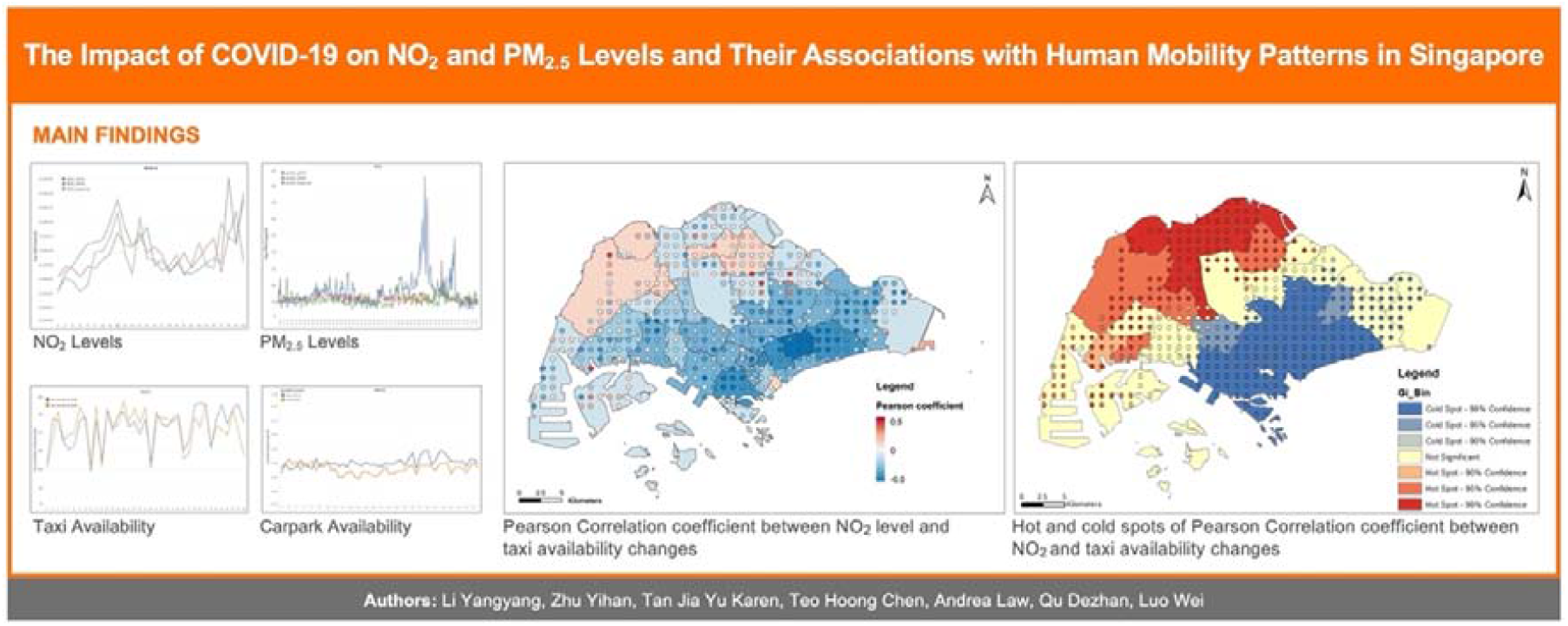

## 1. Introduction

The outbreak of the novel coronavirus SARS-CoV-2 (COVID-19) pandemic has led to unprecedented scales of city-wide and nation-wide lockdowns around the world to reduce human contact and transmissions (Lai et al., 2020; Yin et al., 2021). Resultingly, the restrictions on human mobility within the country drastically decreased emissions coming from public and private vehicular transportations (Doumbia et al., 2021). Singapore, a Southeast Asian megacity, is no exception to this disruption (Dickens et al., 2020; Li and Tartarini, 2020). On 7 April 2020, the Singapore government announced the commencement of a nation-wide lockdown locally known as the “Circuit Breaker” (Ministry of Health, 2020a). This measure heavily restricted outdoor movements including commute to work, school and other activities. This has reportedly led to a reduction in traffic volume by 60% (Tan, 2020) four months into the Circuit Breaker, accounting for at least 44% reduction in emissions from transport-related sources (Jiang et al., 2021). The Circuit Breaker was then followed by three reopening phases (Ministry of Health, 2020b) which led to a rebound of vehicular greenhouse gas emissions as mobility increases (Di Domenico et al., 2020; Zhu et al., 2020; Jiang et al., 2021; Velasco, 2021).

Unlike cities suffering from severe air pollution where lockdown has reportedly resulted in an improvement in air quality (Liu et al., 2021; Bao and Zhang, 2020; Fu et al., 2020; Kerimray et al., 2020), Singapore generally experiences a low level of air pollution mainly in the form of PM_2.5_ even in the pre-COVID days (Zhu et al., 2020). Industries and vehicular emissions constitute one of the constant and major sources of air pollution in Singapore, with transboundary haze periodically affecting Singapore’s air quality particularly in the months of August to October during the Southwest monsoon season (National Environment Agency, 2021). In this context, the change in human mobility in the forms of vehicular traffic during and after Singapore’s Circuit Breaker could therefore potentially lead to a spatial-temporal variation in air quality.

Since the onset of COVID-19 and enforcement of lockdown measures, a plethora of studies has been performed to investigate the effect of lockdown on air pollutions around the world. At regional scale, most of the studies was conducted on Asia (65%), followed by the European Union (18%), North America (6%), South America (5%), and Africa (3%) (Addas & Maghrabi, 2021). At national scale, India is the most studied country (29%), while about 23% of the studies were performed on China and the rest on US (5%), UK (4%) and Italy (Addas & Maghrabi., 2021). These studies generally concluded a substantial reduction of air pollution level and a significant improvement in air quality compared during the COVID-19 lockdown period compared to the pre-lockdown period. A number of air pollutants were being studies: PM_2.5_, PM_10_, NO_2_, NO_x_, O_3,_ SO_2_, CO, NH_3_ etc., among which NO_2_ and PM_2.5_ were found to have the most prominent decline during the lockdown period (Faridi et al., 2021). For instance, Sale city (Morocco) experienced as high as 96% reduction in NO_2_ during the lockdown period (Otamani et al., 2020), and reduction in PM_2.5_ was reported ranging from 76.5% in Malaysia, 58% in Spain and 53.1% in Dehli (Faridi et al., 2021). However, there exist difficulties to compare these results as different methods were used to investigate changes in air pollutants level. Most studies confined the segments of their study periods to only a few months, either comparing the air pollution level during and right before the lockdown period in 2020 or comparing the air pollution level of lockdown period with those in the same period of previous years. Moreover, lockdown measures had been implemented in multiple phases, with various level of restrictions in different countries (Faridi et al., 2021). Some countries enforced complete lockdown, while some implemented partial measures. Such differences led to great uncertainties when comparing air pollution reduction across different countries (Faridi et al., 2021).

The observed decline in air pollutant level has been attributed to the reduction of transportation emissions and restriction on industrial, economic and production activities (Addas & Maghrabi, 2021; Faridi et al., 2021). This is potentially due to that vehicular emissions and road transport are the most notable sources of ambient PM_2.5_ and NO_2_ (European Environmental Agency, 2019). Figure 1 shows that road transport is the largest source of NO_x_ (Including NO_2_) and also, the second largest source of PM_2.5_ (European Environmental Agency, 2019). There is a consensus among researchers that the decrease in PM_2.5_ and NO_2_ levels during lockdown could be largely due to the restricted mobility. However, only few have continued to quantify the effect of lockdown on mobility patterns and their association with air pollution levels. As one of such few studies, Bao and Zhang (2020) reported strong associations between decrease in air pollution and travel restrictions in 44 cities in northern China. In the U.S, Archer et al. (2020) also found strong correlation between decline in NO_2_ level and reduced mobility index (MI), slight increase in PM_2.5_ level, as well as no correlation between PM_2.5_ and MI during April 2020. This was likely due to the unchanged operation of the major sources of PM_2.5_ in the U.S, that is, diesel-based commercial trucks and coal-based electricity generation. Overall, this suggests that the degree of the impact of restricted mobility on air pollution is context and location specific. In Singapore, only a few studies (Li & Tartarini, 2020; Zhu et al., 2020; Velasco, 2021) investigated the impact of circuit breaker on mobility and air pollution. Zhu et al. (2020) concluded that workplace mobility had a significant positive relationship with PM_2.5_ levels between 15^th^ Feb to 1^st^ June 2020. Li and Tartarini (2020) reported a 29% and 45% decrease in PM_2.5_ and NO_2_ levels during 2020 lockdown period (7^th^ Apr – 11^th^ May) compared to the same period in previous years. Such changes of PM2.5 and NO2 were found to be significantly associated with reduced mobility (i.e., HDB carpark availability, Apple driving mobility index, and Google Public transit index) (Li and Tartarini, 2020).

**Figure 1.**
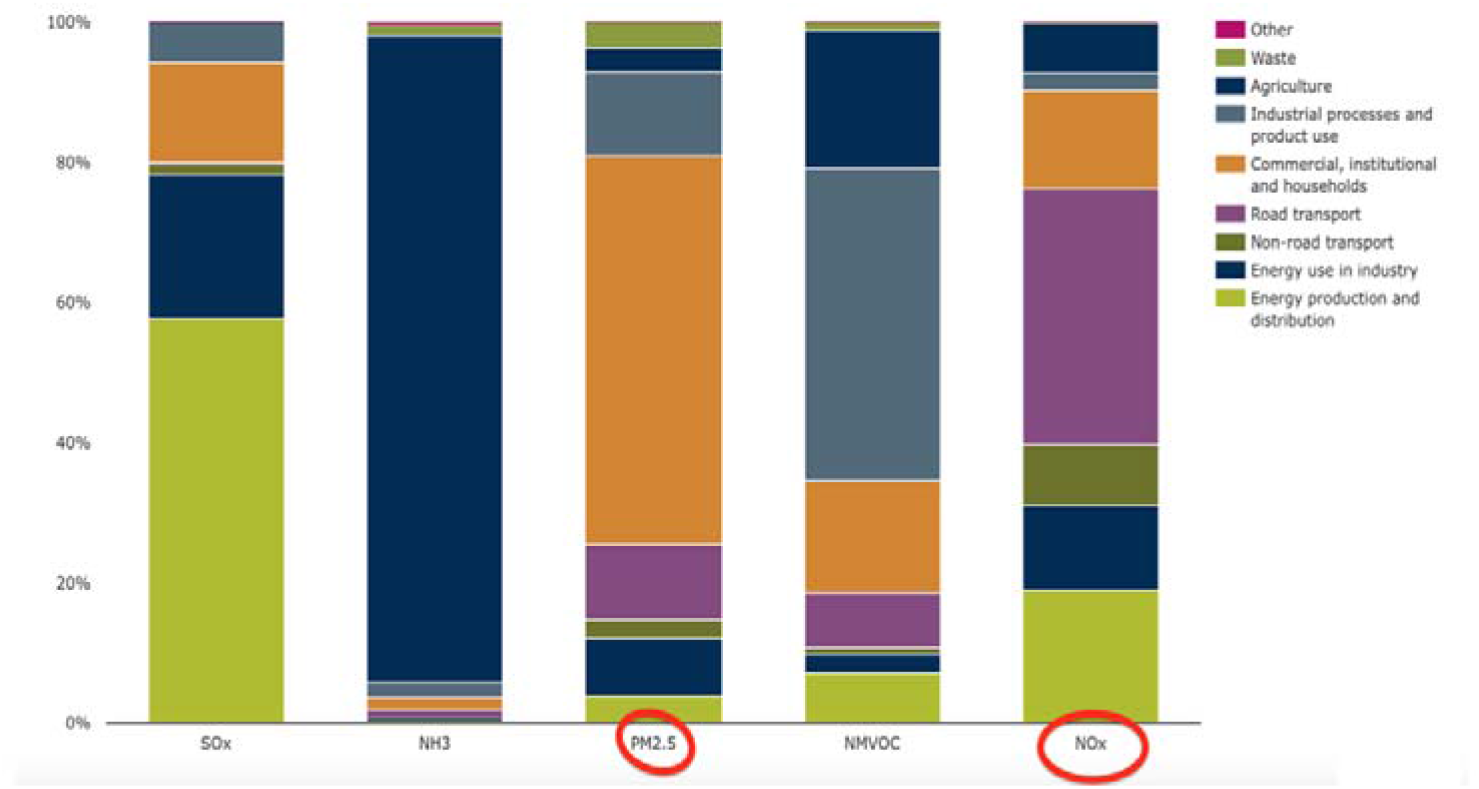
The emission sources of the main air pollutants by sector (modified from European Environmental Agency, 2019).

Besides reduced mobility, it’s well evident that meteorological factors, such as wind speed, wind, wind direction, temperature, precipitation, humidity affect air pollution level (Seinfeld and Pandis, 2016; Yousefian et al., 2020; Jiang et al., 2021; Hua et al., 2021). However, many existing studies did not quantify the effect of meteorological changes on air pollutant level during COVID-19. To date, most studies only compared the air pollution during lockdown with a baseline value (i.e., the estimated 2020 air pollutant level without COVID-19 measures) calculated from historical pollution data. When meteorology was accounted for, the meteorological data were simply used for qualitative interpretation of the measured air pollutant level in 2020 (Faridi et al., 2021). Likewise, Li and Tartarini (2020) only used meteorological data to demonstrate that the monthly averaged meteorological parameters (i.e., temperature, relative humidity, daily rainfall, wind direction) in Singapore during Apr-May 2020 were not significantly different from those of the same period in previous years, thereby justifying their conclusion that the meteorology condition didn’t have much influence on the air pollution level of their period of interest. However, this conclusion may not be reliable as it rests on comparison of monthly average of the studied period alone. This paper thus, argues that the effect of COVID-19 on air quality could potentially be over- or under-estimated. Otmani et al. (2020) observed increased wind speed, humidity and precipitation during lockdown in Sale city (Morrocco), which could decrease air pollutant concentration alongside the positive impact of lockdown restriction and reduced transportation. At the same time, unfavourable meteorological condition can offset the positive impacts of lockdown. Hua et al (2021) reported that the control measures reduced PM_2.5_ by 12 μg/m^3^ in Beijing, whereas the meteorology contributed to an increase of 30 μg/m^3^ in PM2.5, resulting in increased PM2.5 level during lockdown. Therefore, it is important to quantitatively differentiate the impacts of lockdown and meteorology on air pollutant level during COVID-19.

Another research gap is that existing studies mostly assessed the temporal correlation between air pollution level and mobility pattern (Li & Tartarini, 2020; Zhu et al., 2020; Bao & Zhang, 2020; Otmani et al., 2020; Archer et al., 2020). Given both air pollution and mobility are phenomenon with both spatial homogeneity and heterogeneity, this study purports that spatial autocorrelation should be conducted for the correlation value between air pollution change and mobility change during COVID-19. Spatial autocorrelation can help to identify clusters of high correlation between air pollution and mobility, or in other words, areas where mobility variation contributed more to air pollution change.

To account for the variability of the lockdown measures and to increase comparability of results, this study will analyse the air pollutants concentration trend spanning the entire year 2020, rather than confining analysis to a pre-defined lockdown period. Singapore had implemented different phases of measure before opening up in 2020. Expanding period of interest from months to year can avoid bias by covering a wide range of situations from complete lockdown, partial lockdown, gradual opening up, to complete opening up phases. As a result, this study expects the long-term correlation between air pollutants level and mobility indexes to be more reliable than the correlation results calculated solely from the air pollutant and mobility data of the complete lockdown months. To quantitatively differentiate the impact of meteorology from that of lockdown, this study will estimate the baseline air pollution level for 2020 using weather data and haze occurrence records. For instance, a comparatively haze-free condition in Singapore in 2020 (Taufik, 2020) compared to 2019 could suggest a smaller increase in the existing PM_2.5_ and NO_2_ levels. To uncover the spatial variation of the effect of mobility changes on air pollution, this study will perform spatial autocorrelation.

In short, this study aims to 1) quantify the continuous changes in PM_2.5_ and NO_2_ concentration before, during, and after the onset of COVID-19 throughout 2020 in Singapore, 2) to evaluate the association between changes in PM_2.5_ and NO_2_ concentration and the mobility trends throughout 2020, and 3) to identify clusters of stronger correlation between PM_2.5_ or NO_2_ concentration and mobility by performing spatial autocorrelation. Human mobility is commonly understood as the movement of individuals either on foot or via various modes of public and private vehicular transportation. For the purpose of this study, this paper assumed that vehicular transportation operated on private demands such as private cars and taxis closely approximates the degree of human mobility during the study period (Jiang et al., 2021). Results from this study can be used to understand the impact of anthropogenic activities, in particular the traffic patterns, on air quality and to inform effective and targeted strategies for reducing air pollution level in the future beyond the COVID-19 period.

## 2. Material and Methods

This study makes comparisons among two sets of data. The first comparison was conducted at higher temporal resolution (daily) but lower spatial resolution (regional), using PM_2.5_ for air pollution and Housing Development Board (HDB) carpark availability as a proxy for mobility. As approximately 81% of the entire Singaporean population lives in HDB flats (Singapore Statista, 2021), HDB carpark availability can be used as a proxy for human mobility in Singapore. The second comparison was conducted at lower temporal resolution (weekly) but higher spatial resolution (0.01º) data, using NO_2_ for air pollution and taxi availability as a proxy for mobility. Compared to carparks which are stationary and constrained to HDB residential locations, taxies travel around the island and can cover a wider range of locations. Therefore, taxi availability was selected as the mobility proxy for the comparison of higher spatial resolution. A higher carpark availability and a lower taxi availability implies there is a higher human mobility.

### 2.1. Impact of COVID-19 on NO_2_ and PM_2.5_ levels

NO_2_ data was sourced from the Sentinel-5P TROPOMI level 3 product, a global dataset at high spatial resolution of 0.01º and revisit time of around 1 day. The data was downloaded from the Google Earth Engine Data Catalog (Google Developers, 2021) for analysis. PM_2.5_ data was sourced from the National Environment Agency (NEA) Application Programming Interface (API) (NEA, 2021b). The data is of high velocity (temporal resolution) updated every hour but is relatively coarse with observations provided for five regions (Figure 2) in Singapore. The original data pulled from API was resampled first and the details are summarized in Table 1.

**Table 1.** A summary of temporal and spatial resolutions before and after data resampling.

|  |  | PM<sub>2.5</sub> | NO<sub>2</sub> | Weather |  |
| --- | --- | --- | --- | --- | --- |
| Temporal resolution | Original | Hourly | Daily | Up to 1 min interval |  |
|  | Resampled | Daily | Weekly | Daily | Weekly |
| Spatial resolution | Original | Regional | 1.11 km | Station locations |  |
|  | Resampled |  | 0.01° | Regional | 0.01° |

(Notes: The daily and regional weather data was used for 2020 PM<sub>2.5</sub> baseline prediction, while weather data of weekly resolution and 0.01° spatial resolution was used for 2020 NO<sub>2</sub> baseline prediction)

**Figure 2.**
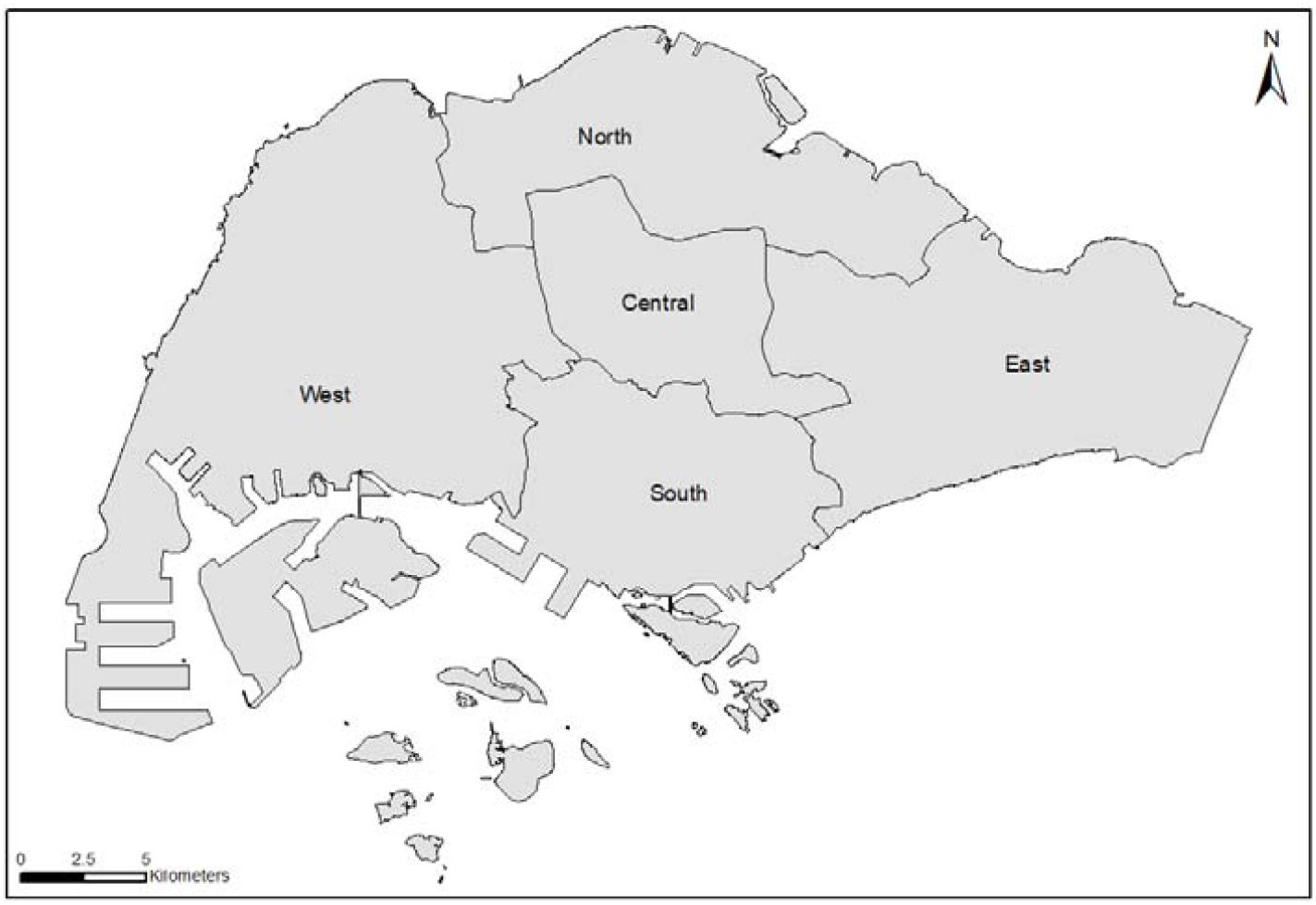
Five regions of Singapore used in this study.

To study how NO_2_ and PM_2.5_ levels changed from 2019 to 2020 as a result of COVID-19, the 2020 air pollutants level without COVID-19 measures were estimated and used as the basis of comparison, which will be referred to as the baseline value in this paper. Changes in NO_2_ and PM_2.5_ levels were calculated from the differences between the measured air pollutant level in 2020 and the baseline. The conventional practice of using the measured NO_2_ and PM_2.5_ in 2019 as the basis of comparison ignores the fact that air quality could be affected by the meteorological condition changes from time to time. For example, a better air quality could be observed with a higher wind speed and precipitation even with a negligible change in mobility (Megaritis et al., 2014; Ahmad et al., 2011). Existing literature have acknowledged that meteorological parameters are important inputs for the predictions of air pollutant levels using machine learning methods (Iskandaryan et al., 2020; Rybarczyk & Zalakeviciute, 2018). Therefore, weather data including rainfall intensity, wind speed, wind direction, temperature, and relative humidity, were used as part of the input variables for NO_2_ and PM_2.5_ baseline prediction. The weather data were retrieved from the NEA API (NEA, 2021c). All weather parameters were first resampled to a daily resolution, where the ordinary kriging method (Wackernagel, 2003) was then used to interpolate them into a spatial resolution of 0.01° from the daily measurements at the locations of weather stations (Figure 3). The interpolated daily weather data was used for PM_2.5_ baseline prediction and resampled further to a weekly resolution for NO_2_ baseline prediction. Moreover, to account for the influence of the haze event in September 2019 on NO_2_ and PM_2.5_ levels, the relative number of haze searches in Google Trends (Google, 2021) from 2019 to 2020 were also used as an input variable. In addition, location information including the longitudes and latitudes, and categorical regions (i.e., Central, North, South, West, and East) encoded using one hot encoder were used as inputs in predicting NO_2_ and PM_2.5_ levels, respectively. The location was considered as input variables as they were related to spatially varied factors such as landcovers and land uses that could influence NO_2_ and PM_2.5_ levels (Xu et al., 2016), or contributions from consistent emissions from sectors other than road transport.

**Figure 3.**
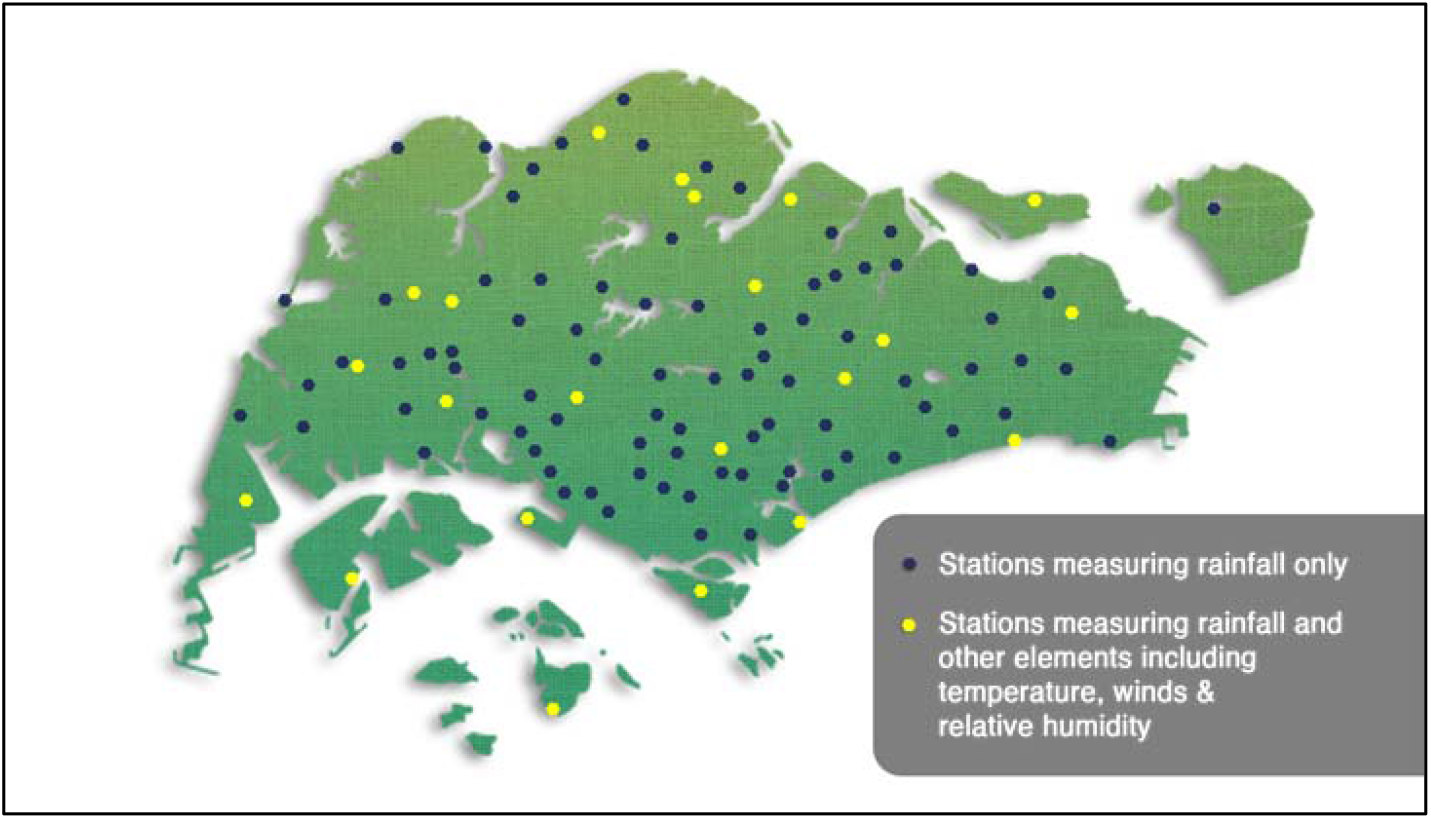
Location of weather stations in Singapore (NEA, 2021d).

In this study, two machine learning methods, Random Forest (RF) and deep neutral network (DNN), were used since they have been commonly used in different studies on air pollutant levels predictions (Zhan et al., 2018; Wang & Song, 2018). The meteorological, location, and haze information in 2019 were input variables and the NO_2_ and PM_2.5_ levels in 2019 were outputs to train and validate each machine learning model. 80% of 2019 data was used in training and the remaining 20% was used to validate the trained model. Then the meteorological, location, and haze information in 2020 were used to predict the baseline NO_2_ and PM_2.5_ levels in 2020. The changes in PM_2.5_ and NO_2_ levels were then calculated from the differences between the measurements in 2020 and the corresponding baseline predictions. The change in PM_2.5_ was obtained in a daily temporal resolution from each region and the change in NO_2_ was obtained in a weekly temporal resolution but with a spatial resolution of 0.01°.

### 2.2. Impact of COVID-19 on taxi availability and carpark availability

HDB Carpark and taxi availability were sourced from the DataMall API (LTA, 2021). Carpark data is available at 1-minute interval, and taxi availability is available at 30 second intervals. Table 2 summarises the original taxi and carpark availability data. The taxi availability data was normalised according to the total taxi amount change in the monthly data of Singapore from 2019 to 2020. The island wide monthly taxi population data was sourced from DataMall (LTA, 2021).

**Table 2.** A summary of temporal and spatial resolutions of carpark and taxi availability before and after data resampling

|  |  | Carpark Availability | Taxi Availability |
| --- | --- | --- | --- |
| Temporal resolution | Original | 1 minute | 30 seconds |
|  | Resampled | Daily | Weekly |
| Spatial resolution | Original | Exact Locations | Exact Locations |
|  | Resampled | Regional | 0.01° |

The changes in taxi and carpark availability were calculated from the differences between the measurements in 2020 and 2019. The change in carpark availability for each region was obtained in a daily temporal resolution at regional scale and the change in taxi availability was obtained in a weekly temporal resolution with a spatial resolution of 0.01º.

### 2.3. Correlations

Two correlation analyses, one between PM_2.5_ level change and carpark availability change, and the other between NO_2_ level change and taxi availability change, were carried out. In order to accommodate the large quantity of data points and the high variety of data properties of these data points, three correlation methods were carried out: Pearson, Spearman’s Rank, and Kendall Rank Correlations (Chok, 2010). Pearson correlation evaluates the linear relationship between two continuous variables. Nevertheless, Pearson correlation is very sensitive to outliers, which may lead to a weak correlation for data distributed in high skewness (Akoglu, 2018; Chok, 2010). Spearman’s Rank Correlation evaluates the monotonic relationship, which is based on the ranked values for each variable rather than the raw data, thus it is not limited to some of the assumptions (e.g., normal distribution of variables). Kendall Rank Correlation is similar to Spearman’s rank correlation but usually has a smaller value (Berg, 2021). Kendall Rank Correlation is calculated based on concordant and discordant pairs, which is less sensitive to errors such as null values in dataset (Tarsitano, 2009). Due to the large quantity of data points (421 locations), it was for the most suitable correlation method. In addition, these data points were geographically distributed, thus their data properties were expected to vary spatially. A single correlation method (e.g., Pearson) may fail to capture other types of bivariate relationships (e.g., non-normally distributed data). Therefore, the correlations between the two sets of variables, as well as their significance, were calculated using three correlation methods: Pearson, Spearman’s Rank, and Kendall Rank Correlations. All the three correlations’ coefficients vary from 1 to -1, indicating positive and negative correlations, respectively. A value close to 0 indicates a very weak correlation.

For PM_2.5_ level and carpark availability, the three correlation methods were conducted both in an island scale and a regional scale. For NO_2_ level and taxi availability, the three correlation methods were performed both in an island scale and at a spatial resolution of 0.01°, with a temporal resolution of every 7 days. Only points with a minimum of 25 observations for both NO_2_ and taxi availability during the studied period were included in the correlation analysis. Correlations were also conducted for each planning area using all observations within the corresponding planning area (Administrative boundaries in Singapore). Planning area boundaries was sourced from Urban Redevelopment Authority (URA, 2021).

### 2.4. Spatial autocorrelation of correlation coefficients

Hot and cold spot analysis was conducted using the Getis-Ord Gi* statistic method to determine statistically significant hot and cold spots for the three correlation coefficients (i.e., Pearson’s r, Spearman’s rho and Kendall’s tau) between NO_2_ and taxi availability changes. To determine the most appropriate threshold distances, the incremental spatial autocorrelation by distance for 10 distance bands was analysed. The distance band with the peak z-score was selected as the threshold distance.

## 3. Results and Discussion

### 3.1. Impact of COVID -19 on NO_2_ and PM_2.5_ levels

The weekly comparison of mean NO_2_ tropospheric column densities in 2019 and 2020 in Figure 4(a) only illustrates weeks with NO_2_ measurements both in 2019 and 2020. Overall, there was a decrease in NO_2_ levels from 2019 to 2020 observed between Weeks 5 to 20. The decrease started from the 5^th^ week, which was before the lockdown started. Similarly, from the daily comparison of regional PM_2.5_ concentrations in 2019 and 2020 in Figure 4(b), an overall decrease in PM_2.5_ levels from 2019 to 2020 was observed. This decrease was observed to be more significant in September 2020, because the severe haze event detected in September 2019 and the haze-free condition in Sep 2020 together contributed to a large difference. However, little difference was found for the regional PM_2.5_ levels due to a limited data spatial resolution.

**Figure 4.**
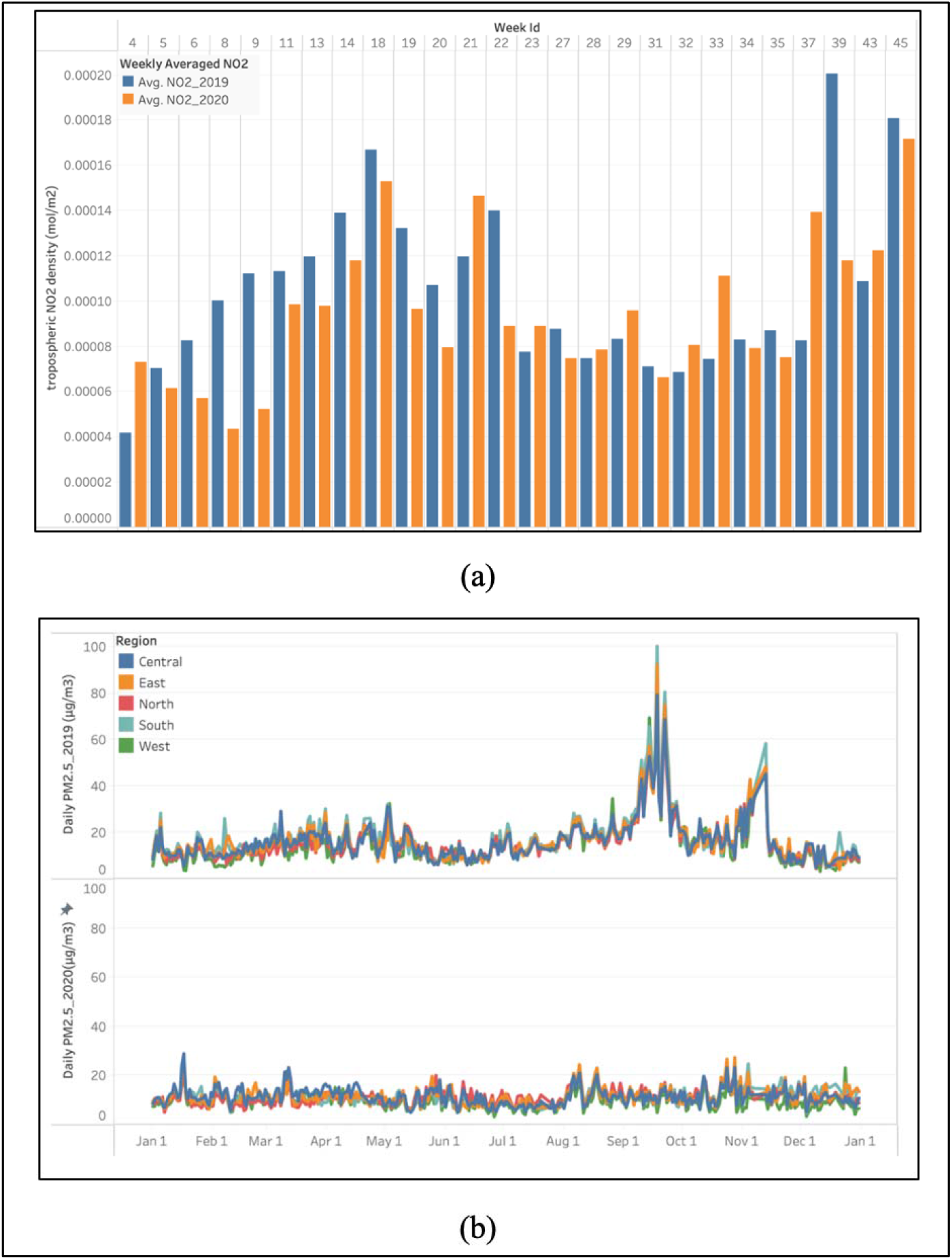
A general comparison of (a) NO_2_ and (b) PM_2.5_ measurements in 2019 and 2020.

Using RF and DNN methods, NO_2_ and PM_2.5_ baselines in 2020 were predicted. Both machine learning models were trained and tested first using 2019 data before baseline predictions. Figure 5 shows the time series data in 2019 that was used in the model training, including NO_2_ meteorological and haze information.

**Figure 5.**
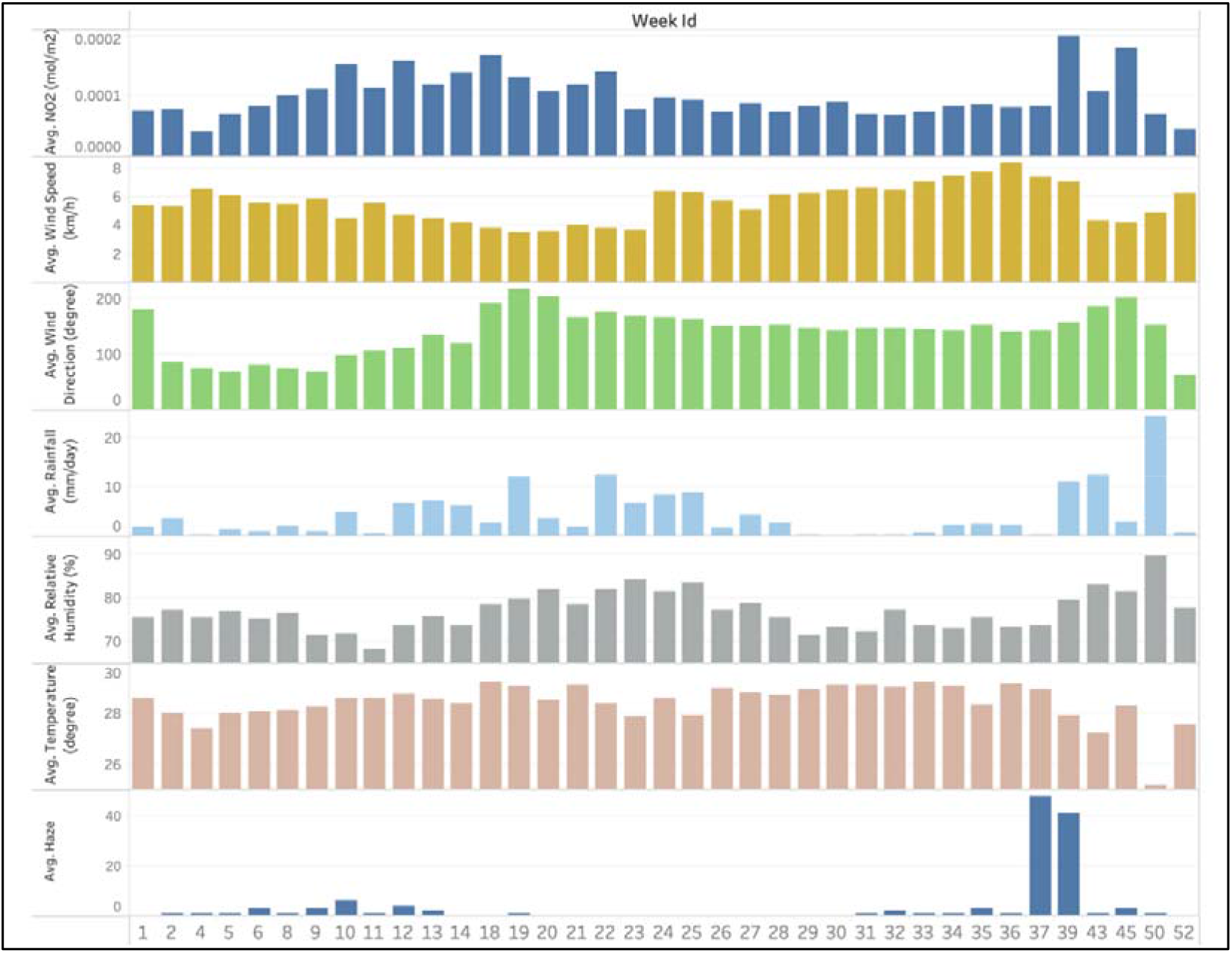
NO_2_, meteorological and haze information in 2019.

Using the two machine learning methods, the comparison of the predicted NO_2_ and the measured NO_2_ levels in 2019 is shown in Figure 6, from training and testing datasets, respectively. The mean absolute error (MAE), the root means square error (RMSE), as well as R^2^ from RF and DNN training and testing datasets are summarized in Table 3. Both methods show similar performance with similar levels of errors (i.e., 5.9e-06 in MAE for RF and 6.0e-06 for DNN, and same RMSE) and accuracy (i.e., 0.971 for RF and 0.964 for DNN). Due to its less computation time, RF was subsequently selected to predict the NO_2_ baseline in 2020, as well as to train the PM_2.5_ model and predict PM_2.5_ baseline in 2020.

**Table 3.**
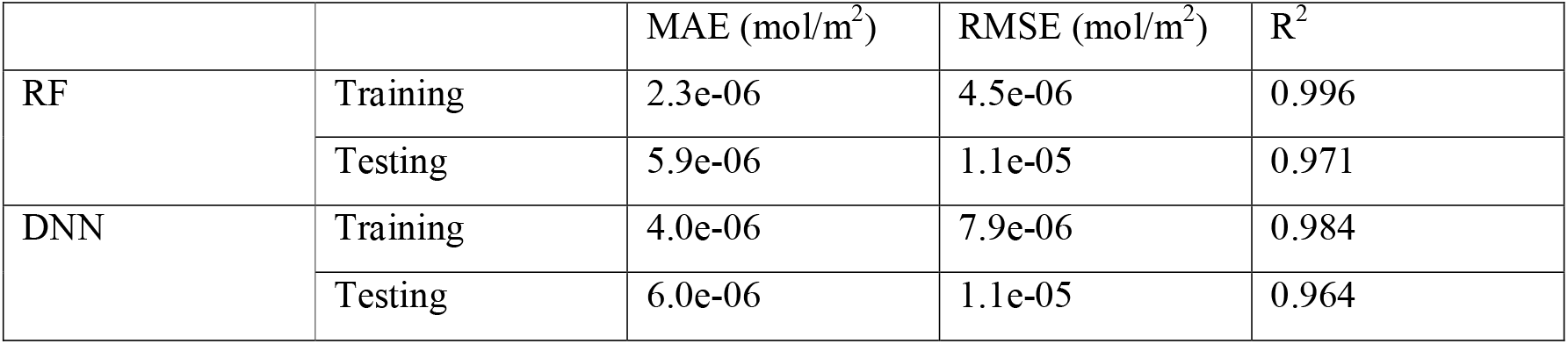
Summarized errors from RF and DNN in NO_2_ modelling.

**Figure 6.**
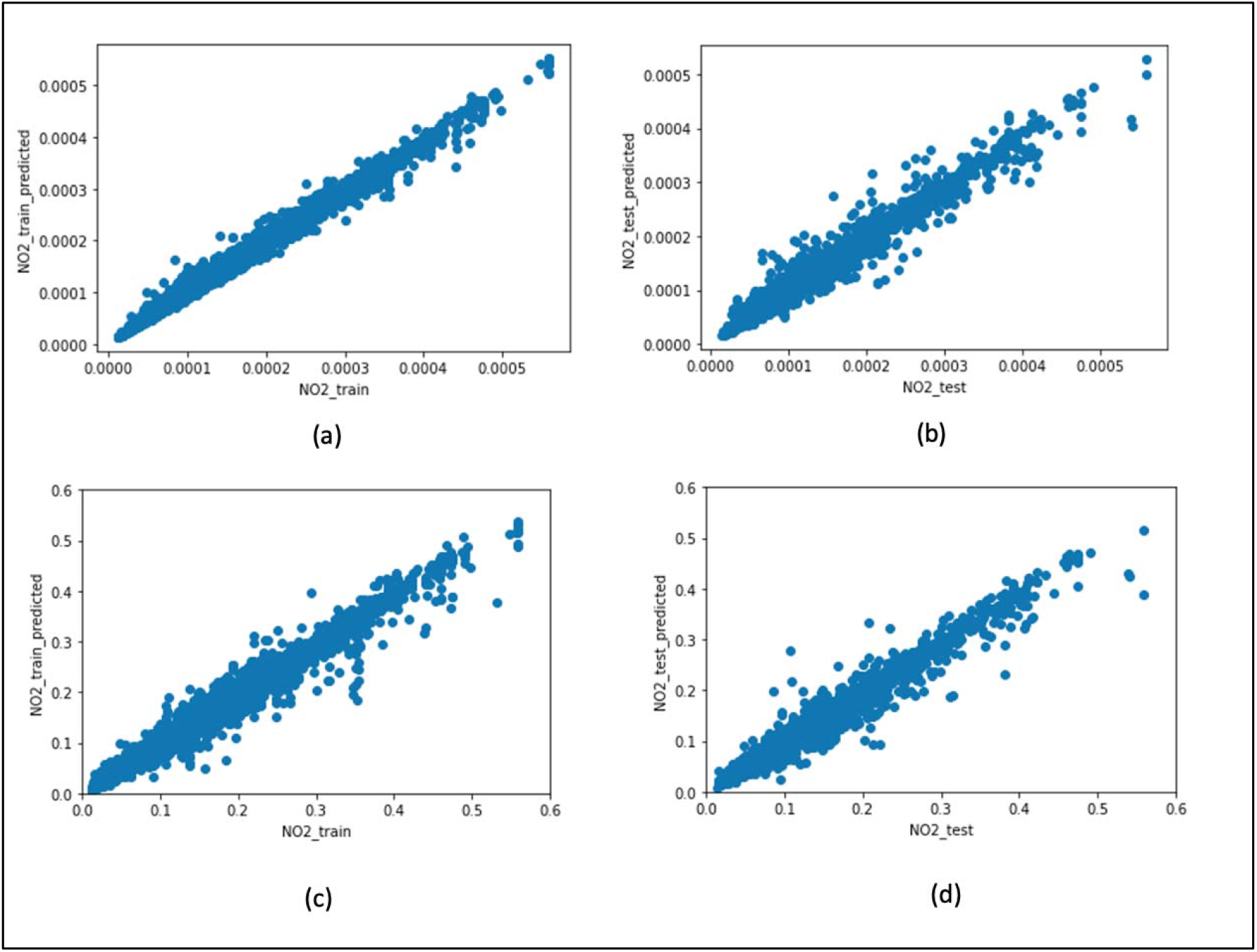
Predicted NO_2_ versus the measured NO_2_ levels (10^−3^ mol/m^2^) from the (a) training and (b) testing datasets using RF and (c) training and (d) testing datasets using DNN.

Figure 7 shows the 2019 PM_2.5_, meteorological and haze time series data used in the model training. Using RF method, the predicted PM_2.5_ and the measured PM_2.5_ levels from the training and testing datasets in 2019 were obtained (Figure 8). The MAE, RMSE and R^2^ in training and testing datasets are summarized in Table 4. The model did not perform well at high PM_2.5_ levels (beyond 60 μg/m^3^), and a difference between predicted and measured values was observed during validation. This could potentially result from the limited number of high PM_2.5_ records (i.e., only during the haze period) for training and testing. However, as there was no severe haze in 2020 and the measured PM_2.5_ did not exceed 60 μg/m^3^. Hence, the predicted results maintain their validity.

**Table 4.**
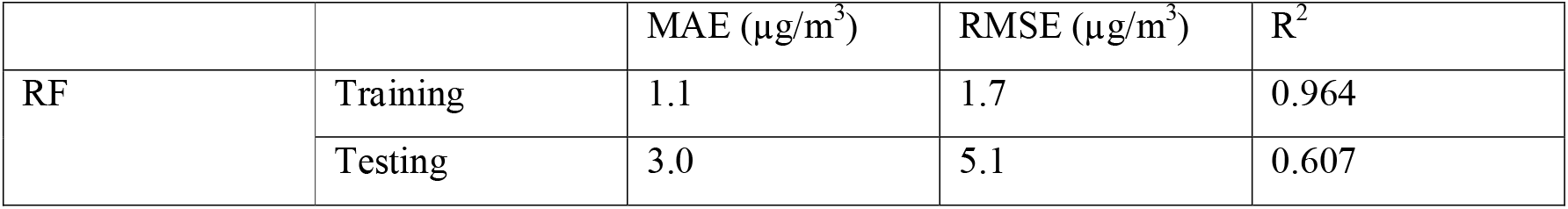
Summarized errors from RF in PM_2.5_ modelling.

**Figure 7.**
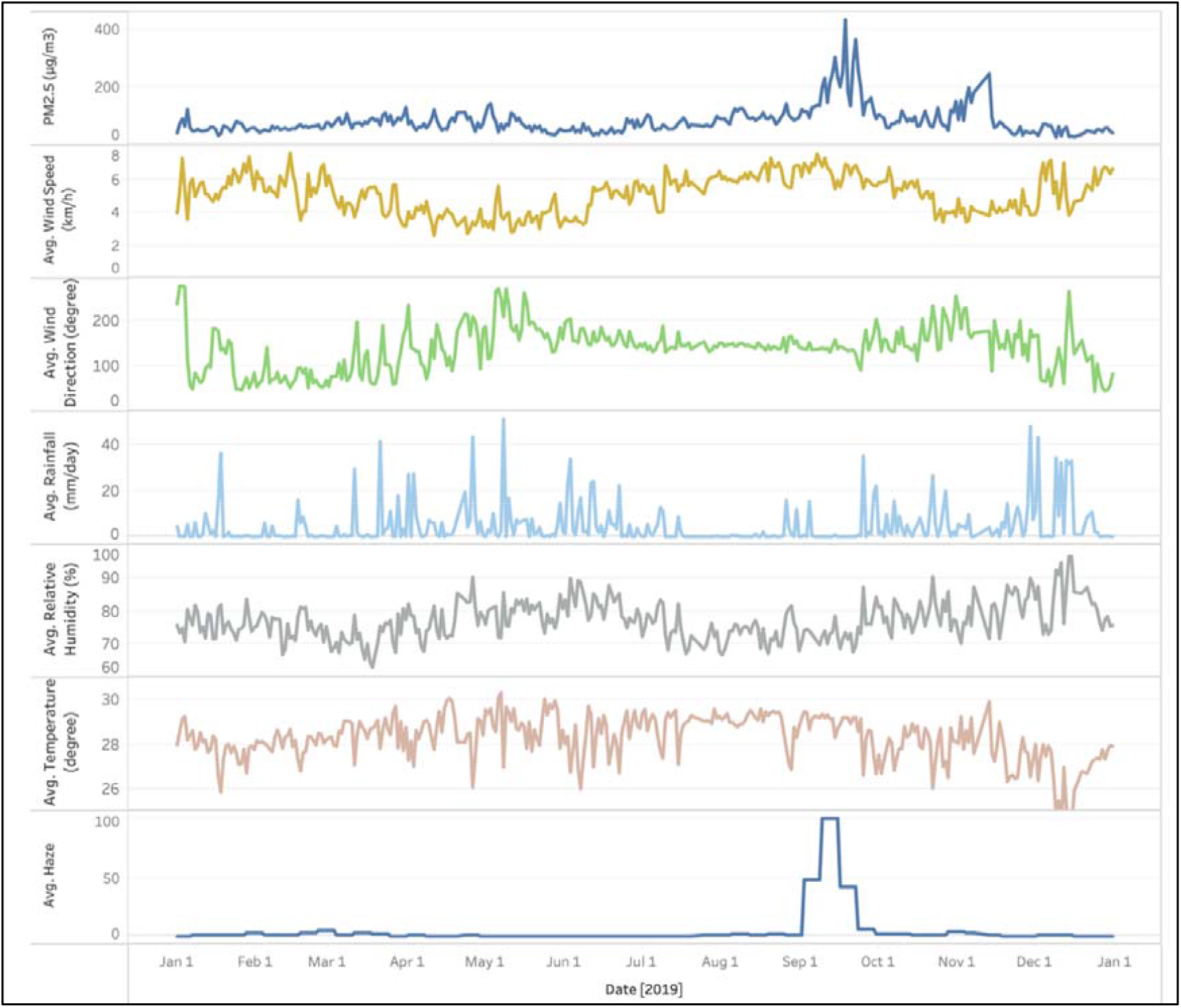
PM_2.5_, meteorological and haze information in 2019.

**Figure 88.**
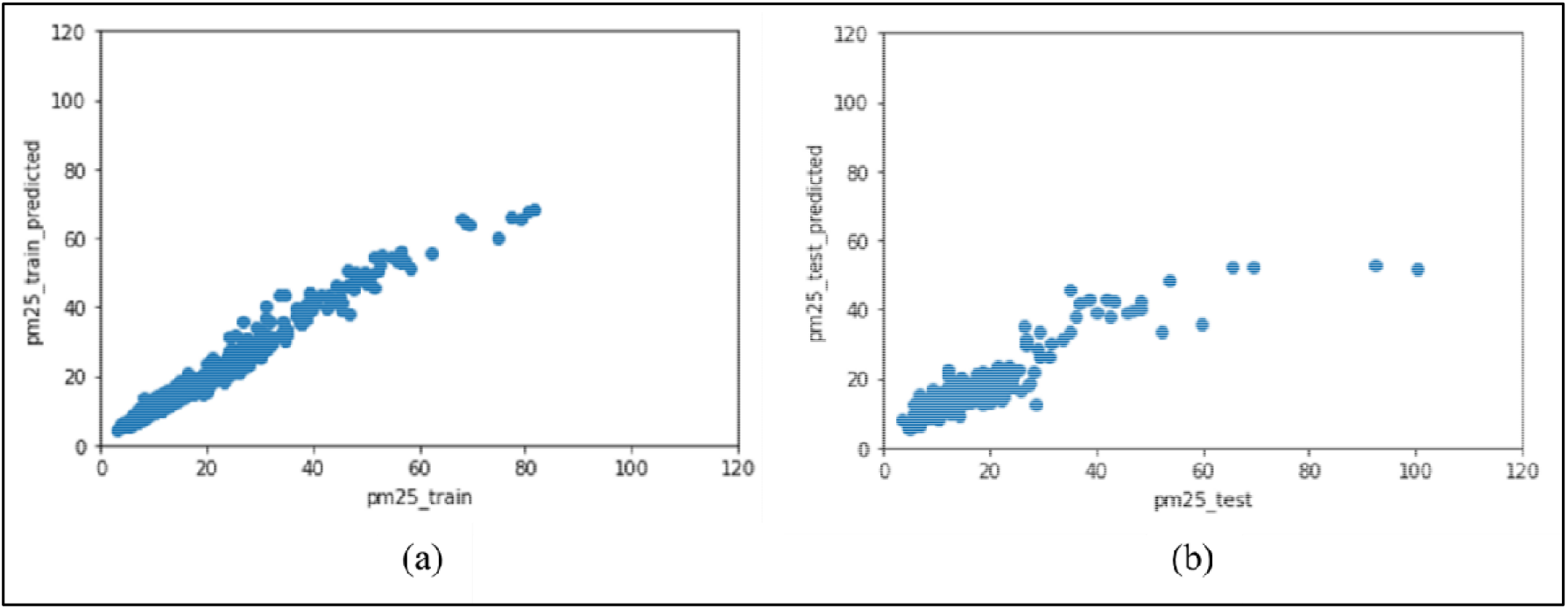
Predicted PM_2.5_ versus the measured PM_2.5_ levels (10^−3^ mol/m^2^) from the (a) training and (b) testing datasets using RF.

From the comparison of feature importance of each input variable in the RF model (Figure 9), a relatively similar degree of importance for NO_2_ modelling while a much higher importance of haze (> 0.5) in PM_2.5_ modelling were found. Locations and wind properties (speed and direction) are comparatively more important (i.e., larger than 0.12) in NO_2_ modelling. Comparatively, the importance of location is very low in PM_2.5_ modelling, implying little difference in the PM_2.5_ comparison across different regions, potentially due to the large study area of the regions preventing finer detection of variation.

**Figure 9.**
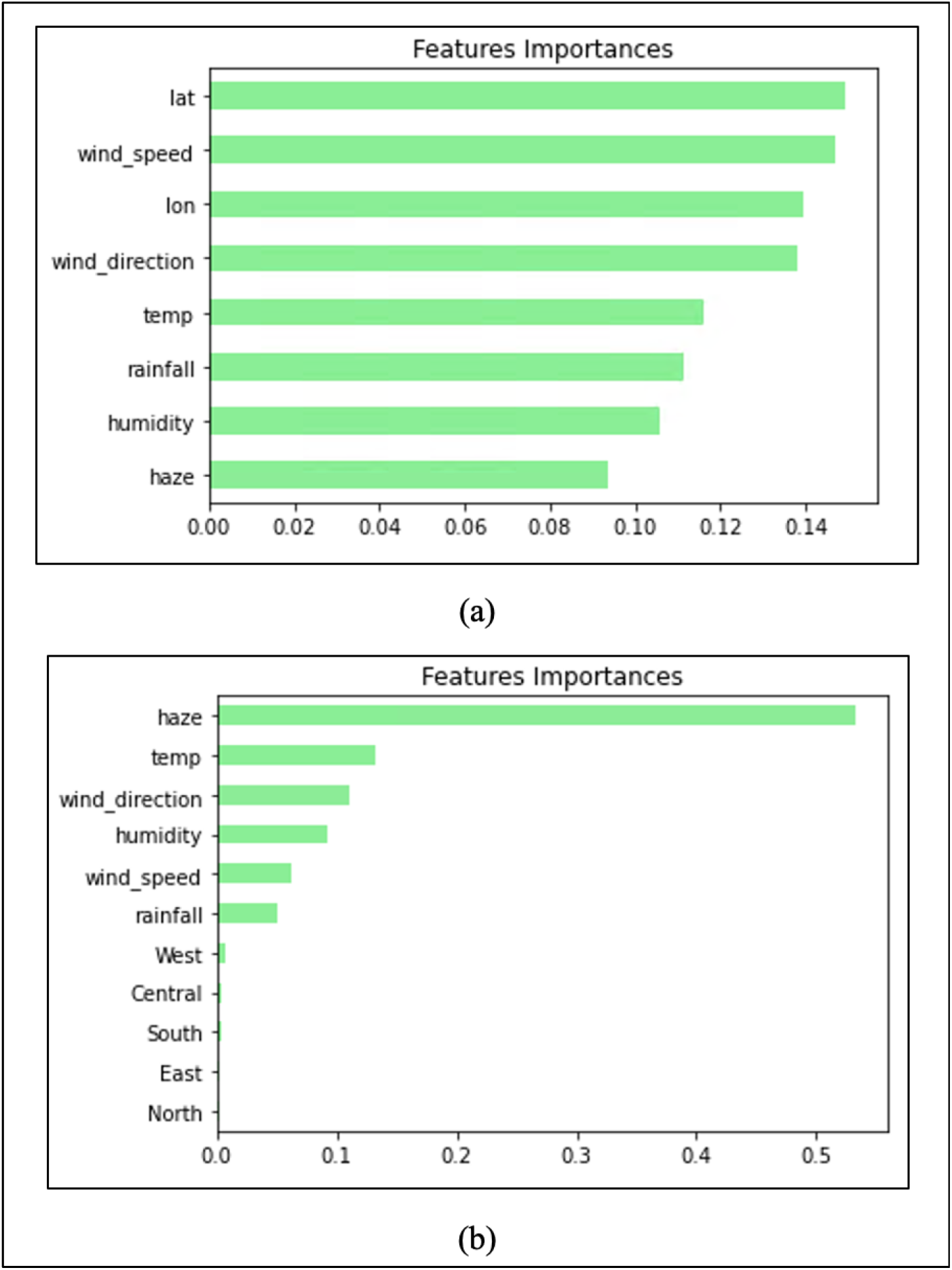
Feature importance in (a) NO_2_ and (b) PM_2.5_ modelling using RF.

From the comparison of the measured NO_2_ levels between 2019 and 2020 in Figure 10 (a), a decrease in NO_2_ levels before Week 20 in 2020 was observed. However, by comparing measured values in 2020 and the predicted baseline use RF method, there was an increase in NO_2_ levels from Week 11 to Week 18, followed by a significant decrease (i.e., about 38%) in NO_2_ levels in Week 20. Similarly, without baseline prediction, the effect of COVID-19 on PM_2.5_ levels could also be overestimated. For instance, in Figure 10 (b), by comparing the measured PM_2.5_ levels in 2019 and 2020, there was a significant decrease in PM_2.5_ levels up to 67% during lockdown period. However, by comparing the predicted baseline and the measurements in 2020, the decrease was only up to 36%. In both NO_2_ and PM_2.5_ baseline prediction, the effect of haze and meteorological parameters in each location was taken into consideration, providing a more reliable estimation on the change in air pollutant levels during COVID-19, including both the lockdown and opening-up period. By only taking lockdown period into study and simply comparing the measured NO_2_ and PM_2.5_ levels in 2020 and 2019, the observed decrease during the lockdown could result in a direct intuition that lockdown helped in reducing NO_2_ and PM_2.5_ levels significantly, which may overestimate the effect of lockdown. However, the overall change in PM_2.5_ is not obvious. This could be due to a change in PM_2.5_ emissions from other sources occurring at the same time when there was a change in PM_2.5_ emissions from road transport. Given household emission is one of the top sources of PM_2.5_ emission (Figure 1), the work-from-home arrangement during COVID-19 can possibly lead to an increased household emission, offsetting the decreased vehicular PM_2.5_ emission on the road.

**Figure 1010.**
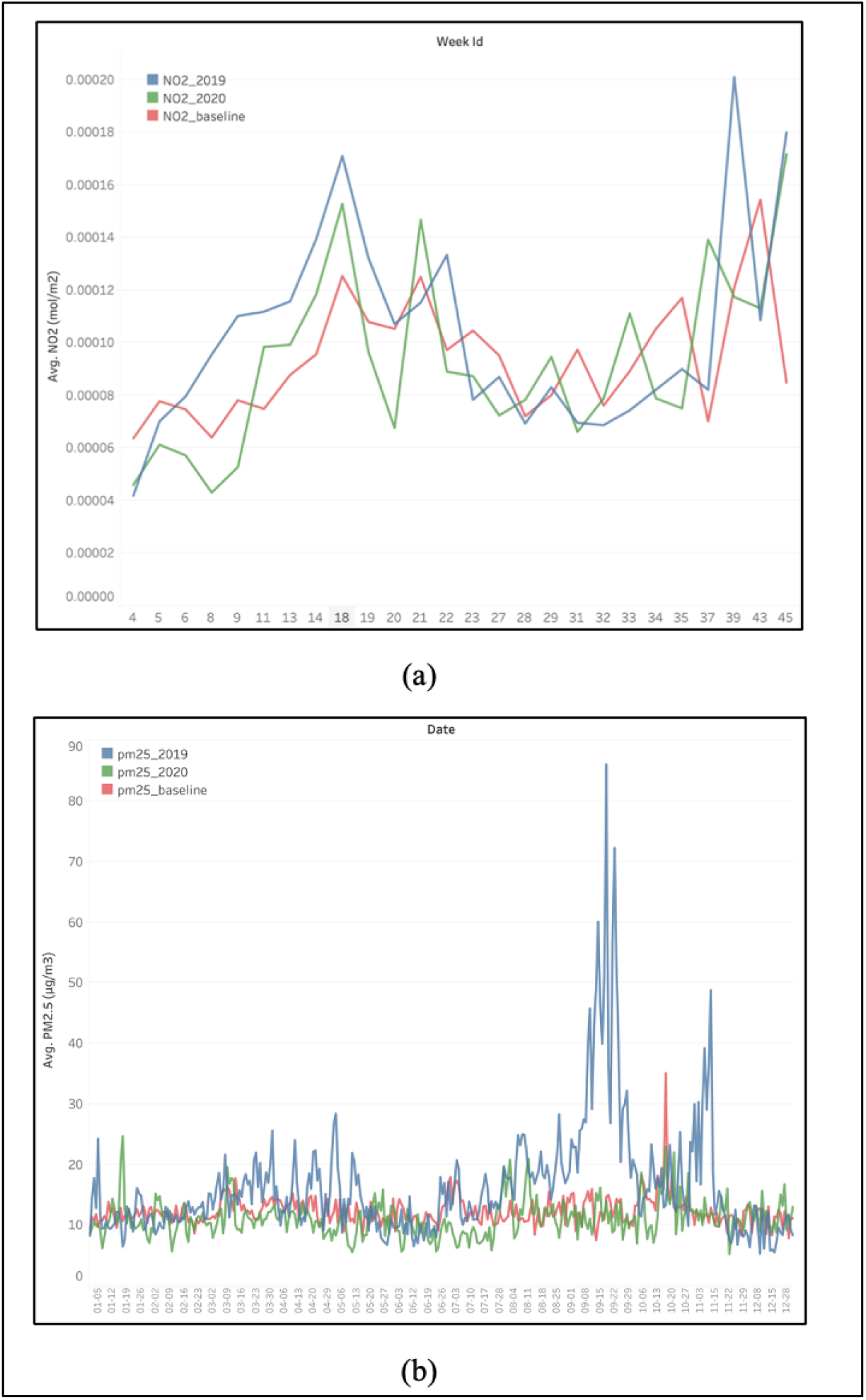
Comparison among measured (a) NO_2_ levels and (b) PM_2.5_ levels in 2019, 2020 and predicted baselines in 2020.

### 3.2. Impact of COVID-19 on carpark and taxi availability

Figure 11 shows the weekly comparison of (a) taxi availability and (b) carpark availability across Singapore in 2019 and 2020. The taxi availability was normalised by the monthly total taxi count. From an overview, across the island, the total number of available taxis has little difference between 2019 and 2020 with no clear pattern. However, it is worth noting that the total number of available taxis in April – June 2020 is higher than the that for the same period in 2019 by up to 12.6%. On the other hand, the weekly comparison of carpark availability across Singapore dropped significantly at the beginning of April (Week 13) in 2020 by up to 9.8% and remained lower than that in 2019 till the end of 2020. In general, pattern of the change in carpark availability was more apparent than that of taxi availability. The sudden drop in carpark availability following the start of lockdown in Singapore in early April suggests that more private vehicles were parked at home and implies a lower mobility. This is congruent with the findings from Li & Tartarini (2020). Similarly, the increase in the number of available taxis during the lockdown in 2020 as compared to 2019 also implies a decrease in human mobility. After the lockdown, carpark availability increased slowly, but remained lower than the same period last year, indicating the continued impact of COVID-19 on mobility. However, taxi availability decreased back to the same level as 2019 in a relatively short period of time.

**Figure 11.**
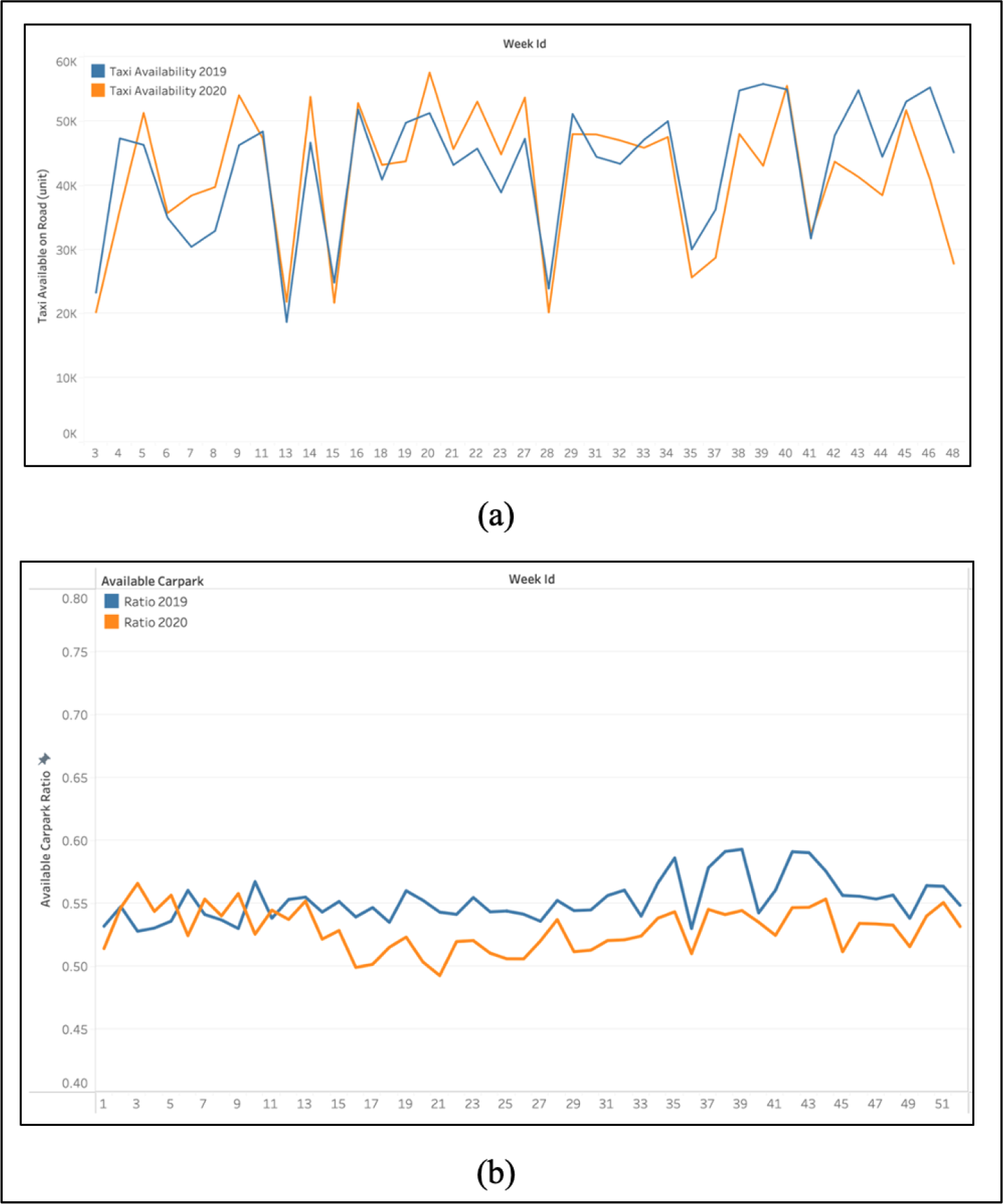
A general comparison of (a) taxi availability and (b) carpark availability in 2019 and 2020

### 3.3. Correlation of PM_2.5_ and carpark availability

Across Singapore, the correlation between PM_2.5_ levels and carpark availability changes is extremely weak and insignificant (Table 5). Among the five regions, the correlation result PM_2.5_ only shows a statistically significant but weak positive correlation in the Central region. This is due to the small change observed in PM_2.5_ levels as discussed in Section 4.1. Moreover, little variation was detected across the five regions mainly due to the data being averaged into five regions, limiting a finer detection of spatial patterns.

**Table 5.** Summary of 3 types of correlation results in between PM_2.5_ level and the average carpark availability changes in the whole island and 5 regions.

| Region | Pearson |  | Spearman's Rank |  | Kendall Rank |  |
| --- | --- | --- | --- | --- | --- | --- |
|  | r | p | rho | p | tau | p |
| **Whole Island** | 0.029 | 0.212 | 0.026 | 0.279 | 0.017 | 0.275 |
| **Central** | 0.122* | 0.021 | 0.103* | 0.049 | 0.071* | 0.046 |
| **East** | 0.077 | 0.149 | 0.093 | 0.079 | 0.062 | 0.079 |
| **North** | -0.052 | 0.331 | -0.057 | 0.282 | -0.039 | 0.266 |
| **South** | 0.043 | 0.413 | 0.032 | 0.547 | 0.023 | 0.521 |
| **West** | 0.078 | 0.139 | 0.063 | 0.237 | 0.043 | 0.228 |

\* Statistics are significant at 95% significance level.

### 3.4. Correlation between NO_2_ and taxi availability

The mean coefficients of Pearson Correlation, Spearman’s Rank Correlation and Kendall Rank Correlation between the correlation of NO_2_ level and taxi availability changes across Singapore are -0.320 (p= 0.049), -0.289 (p= 0.081), -0.210 (p= 0.067), respectively.

In general, the three correlation methods show similar spatial distributions at a spatial resolution of 0.01° (Figures 12). Areas such as the South and East Coast area, show significant negative correlations, suggesting that the change in taxi availability in these places has a stronger correlation with the change in NO_2_ levels when the correlation analysis was performed for each point at a higher spatial resolution. The Pearson correlation coefficients for some points were less than -0.5, exceeding the island-wide average of -0.32, while the correlation coefficients for some points in the northern area were positive, indicating a variation in correlations across the island. The reason for the positive correlation in the north may be due to the inherent limitations of the taxi availability. There are a few points in the north that have very limited number of taxis available (i.e., below 20 units per week), as well as the corresponding change, making the change in taxi availability there may not be representative for the mobility change. This spatial variation may also explain the insignificant correlation results observed in the regional analysis between PM_2.5_ and carpark availability changes.

**Figure 12.**
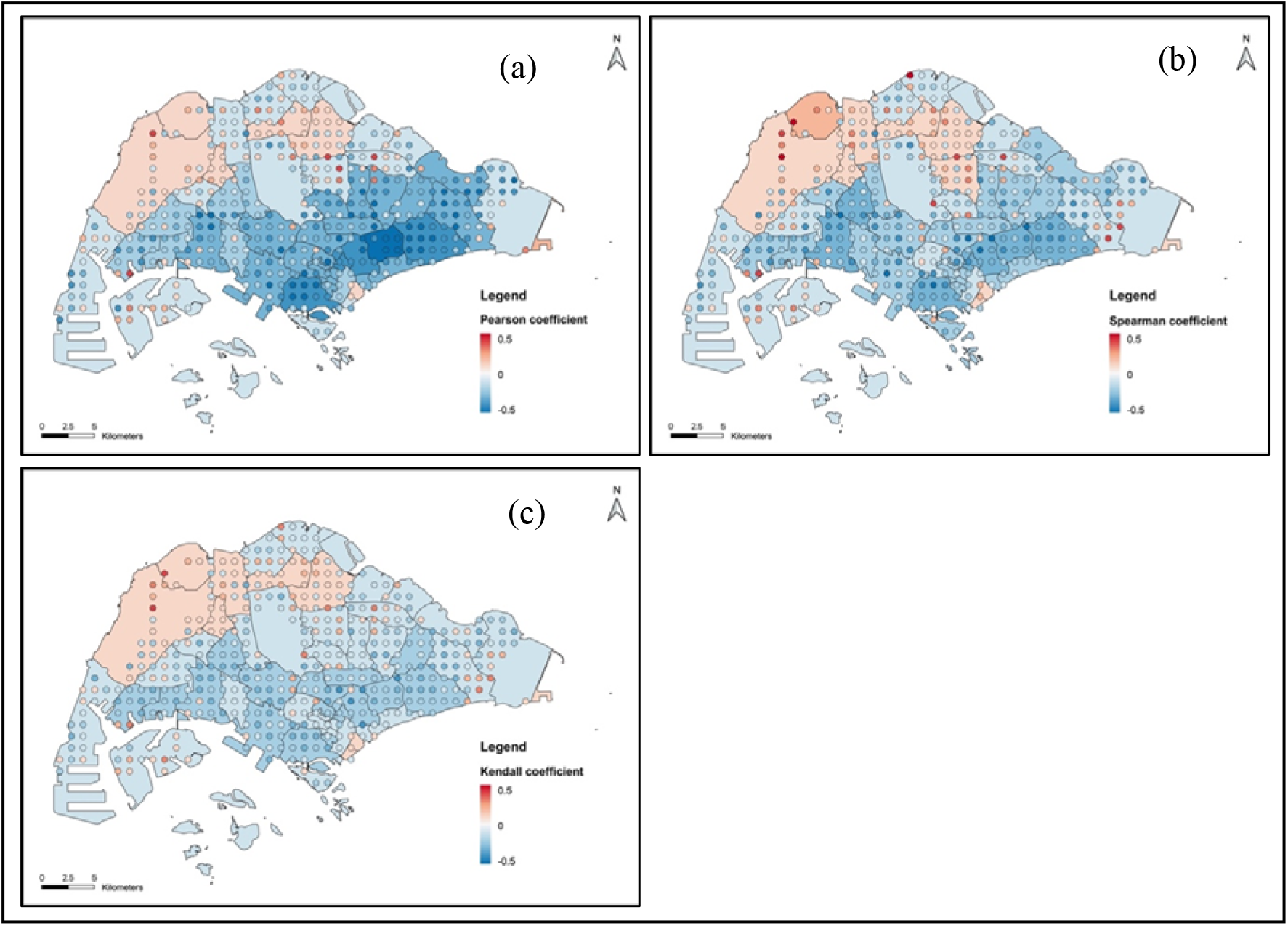
Pearson (a), Spearman’s Rank (b), Kendall Rank (c) Correlation coefficient between NO_2_ level and taxi availability changes in Singapore

### 3.5. Spatial autocorrelation of correlation coefficients

Spatial autocorrelation were run for each NO2 data point, as well as for each planning area using all observations within the corresponding planning area. The hot and cold spot analysis using the three correlation methods (Pearson, Spearman’s Rank and Kendall Rank) revealed generally similar spatial patterns (Figures 13). Notably, there is a clear north-south division in the statistically significant hot and cold spots, with the cold spots in the South and East Coast area, and hot spots in the north. For the non-parametric Spearman’s and Kendall correlations, the strength of the confidence in the south is slightly weaker, but still significant. This suggests that in the South and East Coast areas, a decrease in mobility represented by an increase in taxi availability could more possibly lead to a reduction in NO_2_ due to the cluster of stronger correlations compared to other areas.

**Figure 13.**
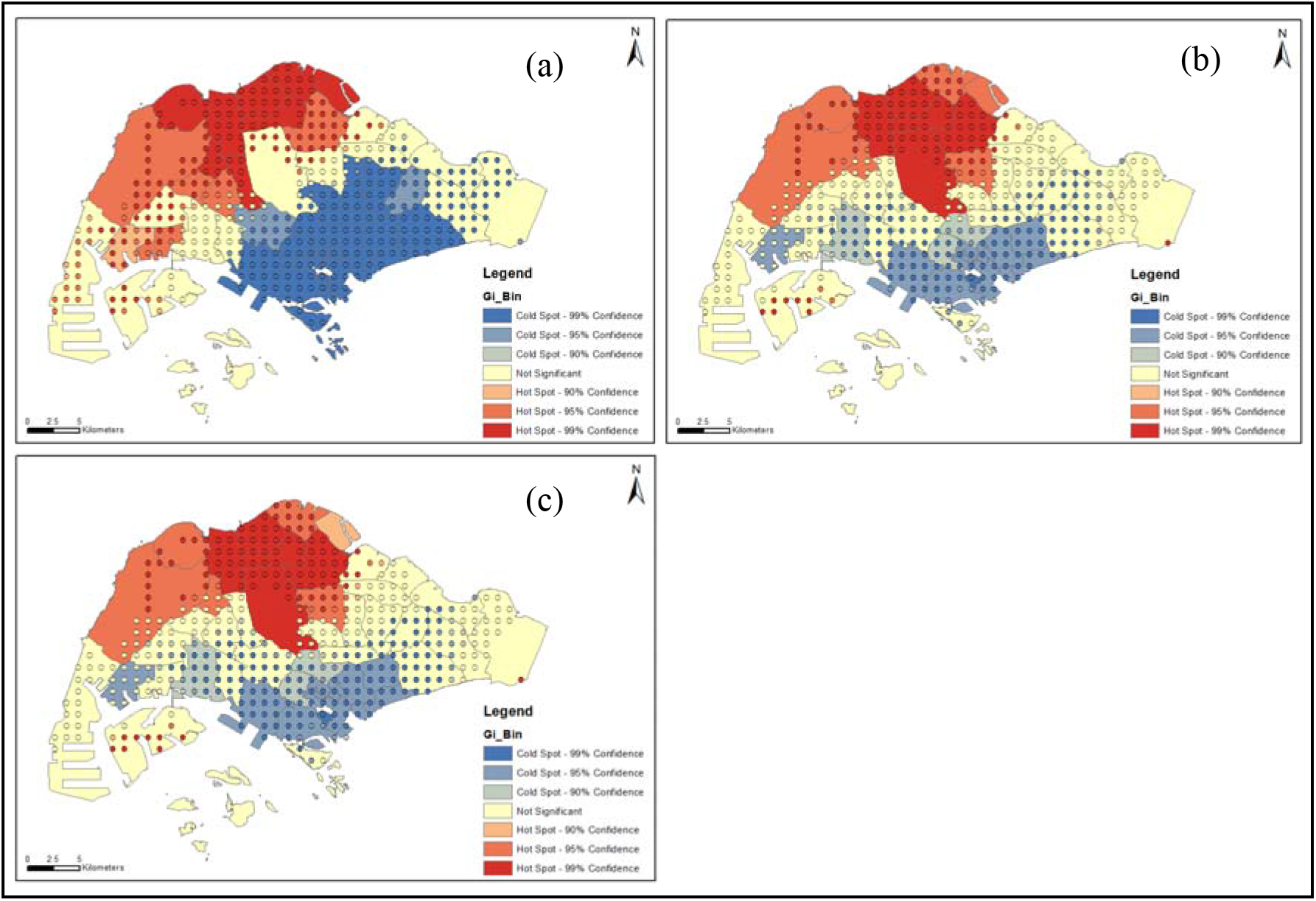
Hot and cold spots of Pearson (a), Spearman’s Rank (b), Kendall Rank (c) Correlation coefficient between NO_2_ level and taxi changes, for each point representing 0.01° pixel, and URA Planning Areas.

## 4. Conclusions

Studies worldwide have observed declines in air pollution due to COVID-19 lockdowns. Similarly, this study also found that in 2020, both NO_2_ and PM_2.5_ declined (by a maximum of 38% and 36%, respectively) from the estimated 2020 baseline in Singapore. However, this decline from the baseline is smaller than the decline from 2019 to 2020. This implies the effect of COVID-19 on reducing NO_2_ and PM_2.5_ levels in Singapore could be over-estimated, if air pollution changes are studied without the baseline prediction taking meteorological factors into consideration. Taxi availability increased and carpark availability decreased by a maximum of 12.6% and 9.8%, respectively, in 2020 from 2019 during lockdown. In general, change in NO_2_ was found more associated with the change in human mobility. Only weak correlations were found between PM_2.5_ levels and carpark availability. However, NO_2_ and taxi availability showed significant correlations and notable spatial patterns of the correlation coefficients were found, especially in the South and East Coast. As such, measures to reduce traffic or vehicular pollution in the South and East Coast could be effective in reducing NO2 levels in these regions. Moreover, we assumed that the change in NO_2_ and PM_2.5_ are mainly due to the change of mobility and the emissions from other sectors are mostly consistent in this study, so future studies can also investigate changes in household or industrial emissions in addition to mobility patterns.

## Data Availability

All data produced in the present study are available upon reasonable request to the authors

## Notes

### Competing Interest Statement

The authors have declared no competing interest.

### Funding Statement

This study did not receive any funding

